# Regression discontinuity design to evaluate the effect of statins on myocardial infarction in electronic health records

**DOI:** 10.1101/2022.04.05.22273474

**Authors:** Michelle C. Odden, Adina Zhang, Neal Jawadekar, Annabel Tan, Adina Zeki Al Hazzouri, Sebastian Calonico

**Author notes:** **Corresponding Author:** Michelle C. Odden, PhD, Associate Professor, Dept. of Epidemiology & Pop. Health, Stanford University School of Medicine, 1701 Page Mill Rd. Palo Alto, CA 94304. sharing senior authorship (Adina Zeki Al Hazzouri, Sebastian Calonico).

## Abstract

Regression discontinuity design (RDD) is a quasi-experimental method intended for causal inference in observational settings. While RDD is gaining popularity in clinical studies, there are limited real-world studies examining the performance of this approach on estimating known trial-established casual effects. The goal of this paper is to estimate the effects of statins on myocardial infarction (MI) using RDD and propensity score matching. For the regression discontinuity analysis, we leveraged a 2008 guideline in the UK that recommends statins if a patient’s 10-year cardiovascular disease (CVD) risk score >20%. We used UK electronic health record data from the Health Improvement Network on 49,242 patients aged 65+ in 2008-2011 (our study baseline) without a history of CVD and no statin use in the year prior to the CVD risk score assessment. Both the regression discontinuity (n=19,432) and the propensity score matched populations (n=24,814) demonstrated good balance of confounders. Using RDD, the adjusted point estimate for statins on MI was in the protective direction and similar to the statin effect observed in clinical trials, although the confidence interval included the null (HR= 0.8, 95%CI: 0.4, 1.4). Conversely, the adjusted estimates using propensity score matching remained in the harmful direction: HR =2.4 (95%CI:2.0, 3.0). Regression discontinuity appeared superior to propensity score matching in replicating the known protective association of statins with MI, although precision was poor. Our findings suggest that, when used appropriately, regression discontinuity can expand the scope of clinical investigations aimed at causal inference by leveraging treatment rules from everyday clinical practice.

## INTRODUCTION

Statins (HMG-CoA reductase inhibitors) are a widely used class of lipid-lowering medications with well-established cardiovascular benefits. A large meta-analysis of randomized, placebo controlled statin treatment trials found a relative risk of 0.76 (95% CI: 0.73, 0.79) for fatal or non-fatal coronary heart disease events.^1^ Statin benefits may be slightly attenuated among older users, as the PROSPER trial among adults 72-80 years found a hazard ratio for myocardial infarction (MI) of 0.81 (95% CI: 0.69, 0.94).^2^ In addition, a recent subgroup analysis of the ALLHAT-LTT trial among adults aged 65 and older reported a hazard ratio of 0.80 (95% CI: 0.62, 1.04) for coronary heart disease events.^3^

In 2008, the National Institute for Health and Care Excellence in the United Kingdom (UK) recommended that statins are indicated for the primary prevention of cardiovascular disease (CVD) if a patient’s 10-year risk for fatal or nonfatal CVD, calculated based on a combination of demographic and clinical factors, is greater than or equal to 20%.^4^ This national-level guideline provides an ideal setting for a regression discontinuity design (RDD) because the treatment is given or withheld only according to the CVD risk score. RDD, an increasingly popular quasi-experimental method, ^5-9^ is aimed at deriving causal estimates from observational data.^10-13^ RDD leverages clinical or policy decision rules in which people fall on either side of a threshold or cutoff for recommending treatment. The assumption is that persons falling just above or below the threshold, as in the 20% CVD risk score cutoff of the UK guideline, are exchangeable, and thus causal estimates can be made near this threshold. This is a distinct approach from propensity score matching, which aims to model the probability of treatment and then match observations based on this probability, thus enhancing exchangeability.^14^ These methods are both of interest in clinical studies where the goal is to estimate treatment benefits and harms, based on observational data. Regression discontinuity and propensity score matching both have the potential to address confounding (e.g. confounding by indication, healthy user bias), yet applied examples directly comparing performance of these two approaches are limited, and thus researchers lack practical guidance for their appropriate implementation.

In the present study, we estimate the effects of statins on myocardial infarction (MI) by a regression discontinuity design and propensity score matching in British adults 65 years and older using a large database of UK electronic health records. By leveraging a national guideline^4^ and a well-established econometric method,^15^ the main goal of this investigation is to evaluate the performance of RDD on replicating known casual effects. Since we know the “truth” regarding the effect of statin on MI from established RCTs,^1^ we use MI as a positive control outcome to calibrate and validate the use of RDD for the study of statin treatment effects more generally. Furthermore, MI is a clearly and objectively defined outcome even in administrative datasets. For contrast, we use motor vehicle accidents (MVA) as a negative control outcome; statin use is not causally associated with this outcome. Finally, we compare the findings from RDD to those from a propensity-score matched analysis and a conventional confounder-adjusted analysis.

## METHODS

### Study Population and Data Source

The IQVIA Medical Research Data, incorporating The Health Improvement Network (THIN), a Cegedim database, contains de-identified electronic health record data from general practitioner practices in the United Kingdom. This research database includes approximately 11 million patients and covers more than 5% of the total population and is representative of the UK general practice population.^16^ A detailed description has been presented elsewhere.^17,18^ THIN data collection began in 2003 and includes a rich collection of variables and diagnoses including demographic information, medical diagnoses (including referrals to specialists), prescriptions, laboratory results, lifestyle factors, measurements taken during medical practice, and free text comments. Data recorded in THIN is very accurate and subject to several assurance procedures.^19-21^ Consultation and prescription rates recorded in the THIN database have been shown to be comparable with published national data estimates.^22^ Furthermore, the validity of the prescription data for epidemiological research has been demonstrated.^19^ In particular, associations between various drugs and outcomes in THIN have shown to be similar, in direction and magnitude, to the associations observed in other UK medical records databases.^19^

### Analytical Sample

Given that the statins guideline was enacted in 2008^4^, we set the baseline period for our study as 2008-2011 to give enough time for the guideline to be implemented. Our initial sample included 76,856 patients aged 65 years and older with an assessment of their CVD risk score recorded anytime during the years 2008-2011. (Figure 1) We then excluded patients who were registered in the THIN database for <6 months prior to their CVD risk score assessment (n=5,133), in order to ensure adequate data on their health history. We also excluded those who had a statin prescription in 2 years prior to their CVD risk score assessment, to mimic the washout period of a randomized controlled trial a new user design. Finally, we excluded persons with a history of CVD prior to their CVD risk score assessment (n=381) as the CVD risk score is intended for persons without a history of events. Our final analytical sample included a total of 49,242 patients.

**Figure 1:**
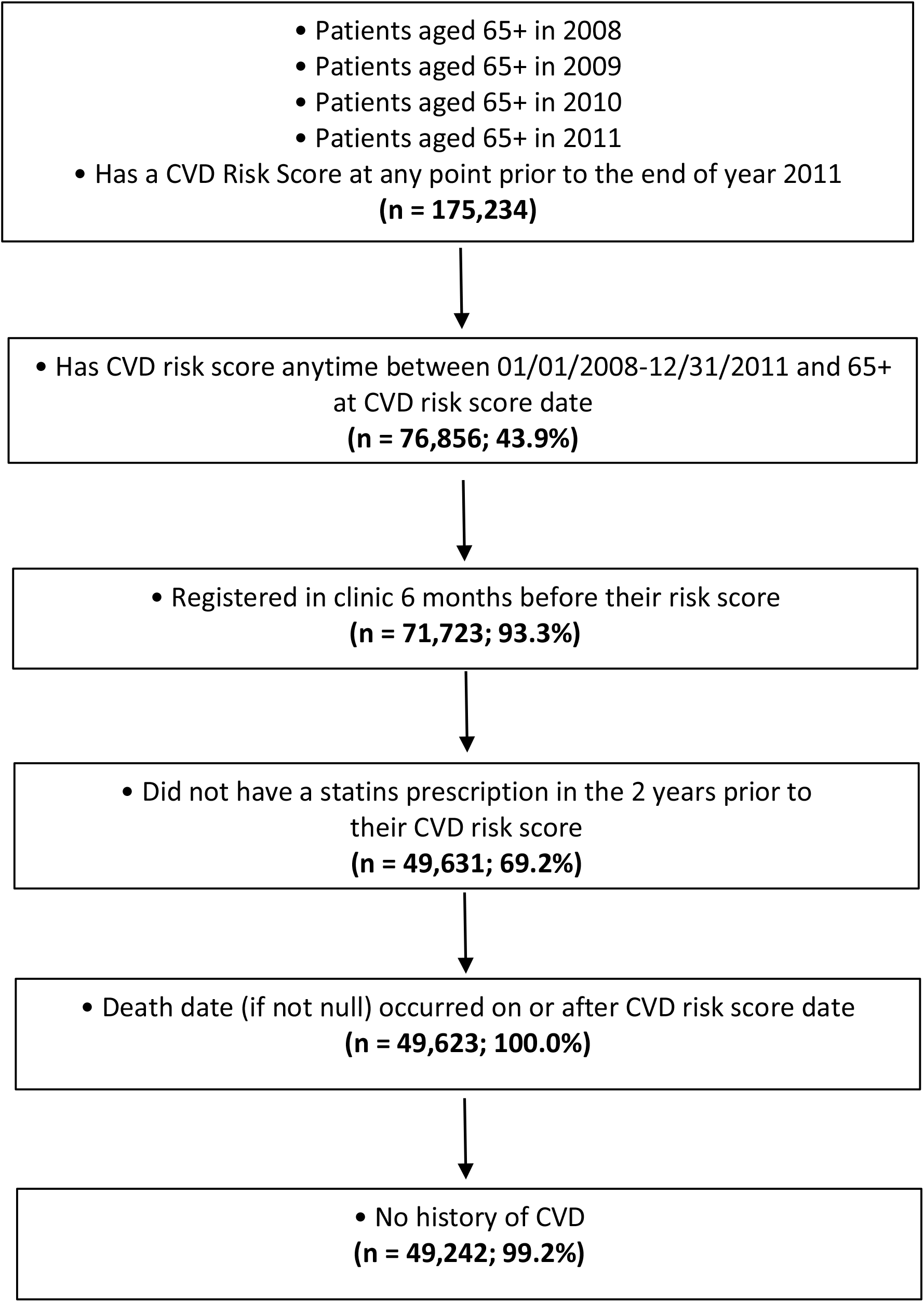
Consort flow diagram for the study analytical sample

#### Assessment of 10-year CVD Risk Score

We used the first CVD risk-score recorded in our study baseline (2008-2011). We included Framingham 10-year CVD risk, QRISK, and QRISK2 scores, which are all prediction algorithms for CVD disease.^23-25^ Of 49,242 participants, 40,259 (81.8%) had CVD risk recorded using the Framingham score. A total of 257 patients had two scores recorded on the same day so we averaged the two. (eTable 1)

#### Assessment of Statin Initiation: Treatment

Statin prescription is recorded in the database using codes from the UK pricing authority,^26^ and its validity has been previously demonstrated.^19^ Statin initiation was defined as a new prescription in a patient with no statin prescription in the prior two years.

#### Assessment of MI and Motor Vehicle Accident: Outcomes

Symptoms and diagnoses are entered using a system of Read codes, a coding system that can be mapped onto ICD-10 codes.^27,28^ The primary outcome of interest was fatal and non-fatal acute myocardial infarction (MI). Events were identified from the list of Read codes provided by IQVIA and cross-checked with members of our study team (MCO, AEM): 323..00, 323Z.00, G30..00, G300.00, G301.00, G301000, G301100, G30..12, G30..13, G30..15, G30..16, G30..17, G301z00, G302.00, G303.00, G304.00, G305.00, G306.00, G307.00, G307000, G307100, G308.00, G309.00, G30B.00, G30X.00, G30X000, G30y.00, 30y100, G30y200, G30yz00, G30z.00, G311000, G311011, G35..00, G350.00, G351.00, G353.00, G35X.00, G360.00, G362.00, G363.00, G364.00, G365.00, G38..00, G380.00, G381.00, G384.00, G38z.00, Gyu3400, Gyu3600. The secondary outcome was injury resulting from motor vehicle accidents, identified from the Medical file using Read codes beginning with T1.

#### Other Measures

Demographic data including age, sex, and marital status. The Townsend score (presented in quintiles) is an area-level measure of deprivation derived from Census data. Additional variables included physical activity (low, moderate, high), smoking status, BMI, diabetes, and hypertension status. For each covariate, we included a category for ‘missing data’. For all covariates, we used baseline values recorded prior to the patient’s CVD risk score.

#### Statistical Analysis

The goal of this analysis was to estimate the effect of statin prescription at baseline (2008-2011) on outcome (MI and MVA) incidence over the next 5 years. To do so, we compared three different modeling approaches: a regression discontinuity model, a traditional multivariable adjusted Cox proportional hazards model, and a propensity score matched Cox proportional hazards model. All models were censored at 5 years of follow-up.

For our regression discontinuity analysis, we estimated the treatment effect of statin use on incident MI using a sharp regression discontinuity for Cox models. RD models can be described by three main components: a score or running variable, a cutoff, and a recommended treatment. We illustrate these concepts in eFigure 1. CVD risk score is the running variable, used to indicate statin treatment. The cutoff value of 20% on the CVD risk score is the threshold for recommending treatment set by the clinical guideline. The *<u>recommended treatment</u>* (e.g., statin treatment vs. no statin treatment) is determined by whether the patient falls above or below the 20% cutoff. In that sense, a sharp regression discontinuity resembles an intent to treat analysis as participants above the cutoff are assumed to have initiated statins, and those below the cutoff are assumed to have *not* initiated statins.^29^ Valid estimation of a causal effect with RD design depends on the *exchangeability* of units below and above the cutoff. The exchangeability concept arises from the intuitive expectation that individuals above and below but “close” to the cutoff should have a similar distribution of *measured* and *unmeasured* prognostic characteristics. The bandwidth, which indicates how far above and below the cutoff to go in order to define our regression discontinuity sample, was chosen based on standard methods balancing the bias and variance of the regression discontinuity estimation,^30,31^ or, equivalently, balancing exchangeability and sample size. We selected a bandwidth of 5.0 percentage points (pp), which means that we included patients with CVD risk scores that are within 5pp of the cutoff (i.e. ranging from 15% to 25%). Furthermore, we explored the sensitivity of the findings to the bandwidth choice. Among those who were eligible for our regression discontinuity experiment (N=49,242), only a total of 19,432 patients were then included in our local regression discontinuity sample determined by the selected bandwidth. Next, using the same bandwidth, we examined covariate balance (i.e. exchangeability) around the 20% cutoff in key confounders including age, sex, marital status, Townsend score, smoking status, physical activity, BMI, diabetes, and hypertension. We first ran unadjusted regression discontinuity Cox models, we then adjusted for potential confounders. Hazard ratios and 95% confidence intervals were calculated using robust standard errors.

Next, we estimated a traditional sex and age-adjusted and multivariable adjusted Cox proportional hazards model. This was conducted in the full sample size of patients who met our eligibility criteria; N=49,242. As such, we examined the relationship of statins initiation in 2008-2011 with incident MI outcomes in the next 5 years, adjusting for the same set of potential confounders as were used in the regression discontinuity model.

Last, we performed a propensity score matching analysis, where we modeled the probability for initiating statins as a function of the same set of confounders. Propensity scores for treatment and control groups were matched one-to-one using the nearest neighbor method. The matched treatment groups were checked for variable balance in their propensity score distributions and absolute mean differences in the matched sample (n=24,814). We then ran a Cox model on the new matched sample evaluating the relationship of statins initiation with MI outcomes, unadjusted and then adjusted for the same confounders.

We explored injuries from motor vehicle accidents (MVA) as a negative control outcome, and used a 10-year follow-up (instead of 5 year as in MI outcome) to account for the lower event rate. We evaluated the relationship between statin use and injuries from MVA using unadjusted and adjusted regression discontinuity and traditional Cox models. The adjusted models did not converge with the inclusion of diabetes, so the set of adjustment variables for these models included age, sex, marital status, Townsend score, smoking status, physical activity, BMI, and hypertension.

R version 4.0.3 with tidyverse, rdrobust version 0.99.5,^32^ survival version 3.2-7 and MatchIt packages were used for the analyses.

#### Patient and public involvement

No participants were involved in setting the research question or the outcome measures, nor were they involved in developing plans for recruitment, design, or implementation of the present study/analysis. No participants were asked for advice on interpretation or writing up of results. We recognize that public involvement has great value and contributes to improving the quality of research, but the scope of the present study does not involve patients. All results are disseminated to study participants and a larger audience via a website, which has a Resources Hub (https://www.the-health-improvement-network.com/resources-hub).

## RESULTS

The cohort of 49,242 patients who met our inclusion criteria had a mean age of 69.8 years (SD 4.2) and a little over half were women (56.1%). (**Table 1**) The majority were nonsmokers, and about a third each had normal, obese, and very obese BMI. Nearly half had hypertension (43.2%) and only 2.4% were diabetic. Most patients had 10-year CVD risk assessed by the 1991 Framingham risk score, and the mean score was 23.2% (median = 21.6%). (**eTable 1**) Among the 21,326 participants who had a 10-year CVD risk score below the 20% cutoff, 11.9% initiated statins by the end of 2011. Among the 12,114 patients who had a 10-year CVD risk score at or above the 20% cutoff, 35.4% initiated statins by the end of 2011. (**eTable 2**)

**Table 1:** Baseline characteristics of patients at time of CVD-risk score assessment

| N | Overall<br>N=49,242 |
| --- | --- |
| Age, years (Mean, SD) | 69.8 (4.2) |
| Sex (N, %) |  |
| Male | 21,625 (43.9) |
| Female | 27,617 (56.1) |
| Marital Status* (N, %) |  |
| Partnered | 2,187 (4.4) |
| Single | 9,473 (19.2) |
| Missing | 37,582 (76.3) |
| Smoking Status (N, %) |  |
| Not currently smoking | 42,986 (87.3) |
| Currently smoking | 6,085 (12.4) |
| Missing | 171 (0.3) |
| Physical Activity (N, %) |  |
| Moderate activity or less | 14,292 (29.0) |
| High activity | 2,279 (4.6) |
| Missing | 32,671 (66.3) |
| Townsend Score (N, %) |  |
| Lowest 3 quintiles | 34,436 (69.9) |
| Upper 2 quintiles | 9,794 (19.9) |
| Missing | 5,012 (10.2) |
| BMI Categories (N, %) |  |
| Less than 25 kg/m<sup>2</sup> | 16,550 (33.6) |
| 25-30 kg/m<sup>2</sup> | 19,720 (40.0) |
| 30-35 kg/m<sup>2</sup> | 8,084 (16.4) |
| 35+ kg/m<sup>2</sup> | 3,023 (6.1) |
| Missing | 1,865 (3.8) |
| Hypertension (N, %) |  |
| No | 27,968 (56.8) |
| Yes | 21,274 (43.2) |
| Diabetes (N, %) |  |
| No | 48,042 (97.6) |
| Yes | 1,200 (2.4) |

\*partnered includes married, co-habiting, stable relationship; single includes single, widowed, divorced, separated.
All patient characteristics were measured at the visit prior to when their CVD risk score was assessed.

Potential confounders were balanced using a regression discontinuity design with a bandwidth of 5.0% (n=18,718), and the confidence intervals for the differences above and below the 20% CVD risk score cutoff included the null. (Table 2)

**Table 2:** Balance of key confounders above and below the 20% CVD risk score cutoff, using a regression discontinuity design*

| Variable | N | N <20% \| ≥20%<br>risk | Coefficient<sup>†</sup> | Confidence<br>Interval | p-value |
| --- | --- | --- | --- | --- | --- |
| Age, years | 18,718 | 9,670 \| 9,048 | -0.139 | -0.362, 0.329 | 0.93 |
| Gender<br>(Woman/man) | 18,718 | 9,670 \| 9,048 | -0.030 | -0.082, 0.003 | 0.07 |
| Marital status<br>(Partnered vs Single) | 4,509 | 2,432 \| 2,077 | 0.024 | -0.056, 0.080 | 0.73 |
| Townsend Score<br>(4<sup>th</sup> - 5<sup>th</sup> vs 1<sup>st</sup>-3<sup>rd</sup><br>quintile) | 16,934 | 8,785 \| 8,149 | 0.012 | -0.035, 0.043 | 0.84 |
| Townsend Missing | 18,718 | 9,670 \| 9,048 | 0.018 | 0.002, 0.054 | 0.03 |
| Smoking Status<br>(Current vs. Not<br>current) | 18,666 | 9,646 \| 9,020 | 0.007 | -0.009, 0.035 | 0.24 |
| Physical activity<br>(High vs<br>Low/moderate) | 6,380 | 3,346 \| 3,034 | 0.030 | -0.007, 0.105 | 0.09 |
| BMI kg/m<sup>2</sup> | 18,116 | 9,393 \| 8,723 | -0.087 | -0.413, 0.441 | 0.95 |
| Diabetes | 18,718 | 9,670 \| 9,048 | -0.005 | -0.016, 0.002 | 0.15 |
| Hypertension | 18,718 | 9,670 \| 9,048 | -0.008 | -0.046, 0.042 | 0.92 |

\*This sample was restricted to those within the bandwidth of 5.0
<sup>†</sup>Represents above minus below the cutoff

A total of 12,412 pairs were matched for a matched analysis sample size of 24,814. Before matching, female sex, smoking status, and BMI were most imbalanced among statin users and non-users. After matching, all absolute mean differences in covariates were less than 0.05. (**Figure 2**) Propensity scores ranged from 0.08 to 0.70 before matching and 0.46 to 0.79 after matching (**eFigure 3**).

**Figure 2:**
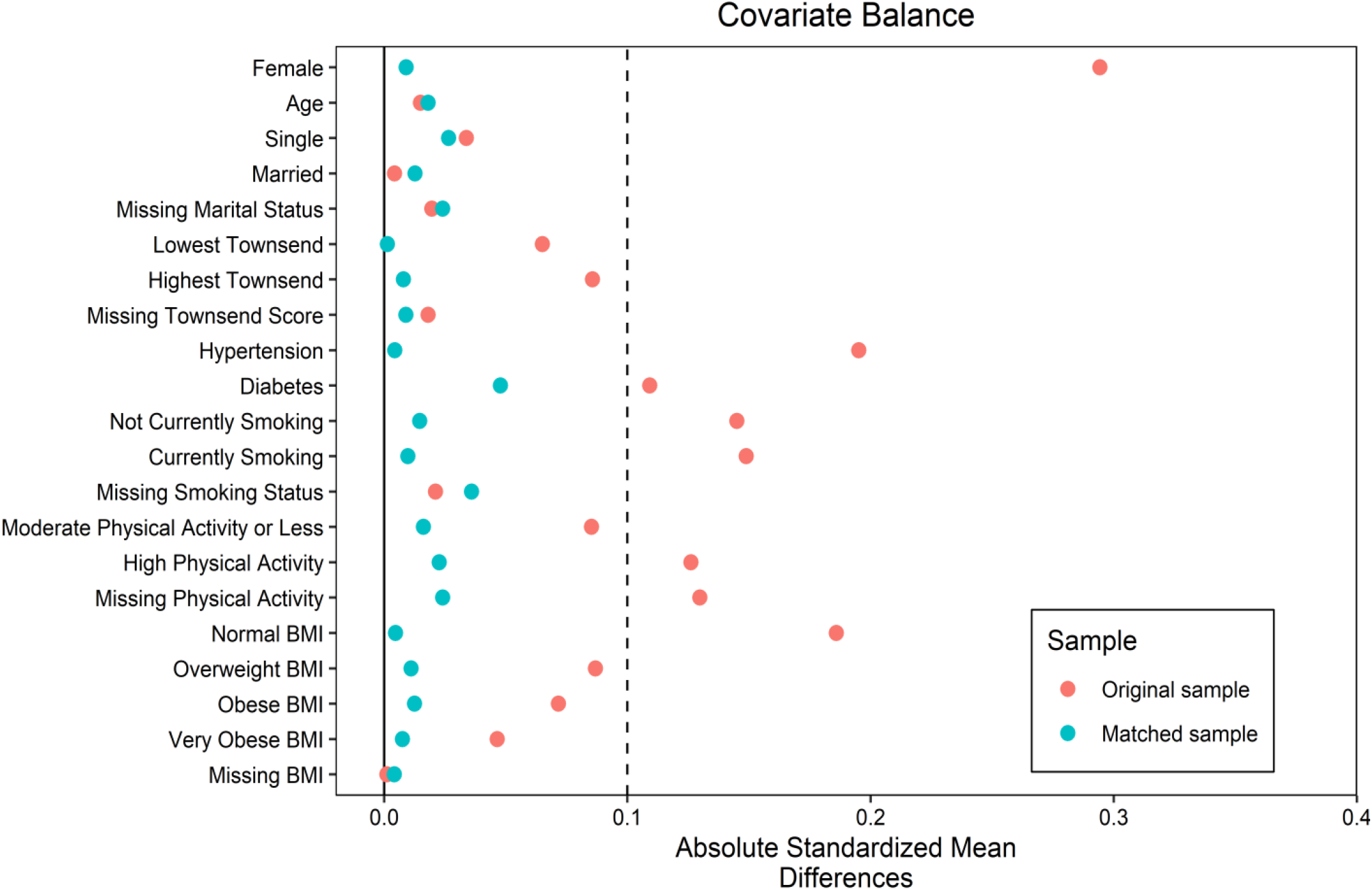
Standardized absolute difference in key confounders before and after matching

Among the 49,242 eligible patients, 575 (1.2%) had an MI over the next 5 years. The algorithm selected a bandwidth of 5.0 and we observed that the HR of MI was robust and relatively the same across a range of bandwidth values (**eFigure2**, bandwidth calibration). Based on a regression discontinuity model, the association between statin treatment and MI was in the protective direction, although the confidence interval included the null; the adjusted regression discontinuity Cox model estimate was 0.79 (95% CI: 0.44, 1.43). (**Table 3**) In contrast, based on traditional Cox models, the estimate was in the harmful direction (adjusted HR = 2.51, 95%CI: 2.12, 2.97). Results from propensity score matching did not meaningfully differ from the standard Cox estimates: the adjusted propensity score matched Cox model HR estimate was 2.41 (95% CI: 1.96-2.99). (**Table 3**)

**Table 3:** A comparison of estimates of the association of statins use (2008-2011) and incident MI over 5 years, based on different analytic approaches

| Model | N | HR | Confidence Interval | p-value |
| --- | --- | --- | --- | --- |
| **Regression Discontinuity Model\*** |  |  |  |  |
| Sex, Age adjusted | 19,432 | 0.80 | 0.44, 1.44 | 0.45 |
| Fully<sup>†</sup> adjusted | 19,432 | 0.79 | 0.44, 1.43 | 0.44 |
| **Traditional Cox Model** |  |  |  |  |
| Sex, Age adjusted | 49,242 | 2.69 | 2.28, 3.17 | <0.001* |
| Fully<sup>†</sup> adjusted | 49,242 | 2.51 | 2.12, 2.97 | <0.001* |
| **Cox Model in Propensity Score Matched Subset** |  |  |  |  |
| Sex, Age adjusted | 24,814 | 2.44 | 1.97, 3.01 | <0.001* |
| Fully<sup>†</sup> adjusted | 24,814 | 2.41 | 1.96, 2.99 | <0.001* |

\*This sample was restricted to those within the bandwidth of 5.0
<sup>†</sup>Adjusted for age, sex, marital status, Townsend score, smoking, physical activity, BMI, hypertension, diabetes.

While our choice of a 5-year follow-up period for MI was based on a priori RCTs, in exploratory analyses, we evaluated the regression discontinuity Cox model at different lengths of follow-up and found a stronger effect at 2 years (0.58, 95% CI: 0.22, 1.57) that attenuated towards the null at 10 years (0.96, 95% CI: 0.59, 1.54). (**eTable 3**)

Among the 49,242 eligible patients, 160 (0.3%) had recorded injuries due to MVA over the next 5 years, and 213 (0.4%) within 10 years. Given the low MVA event rate and the fact that the effect size estimates were less variable using the 10 years vs. the 5 years follow up period, so we selected the 10-year for our MVA analyses. (**eFigure 2**, bandwidth calibration) The estimate for the association between statin treatment and incident MVA from the regression discontinuity Cox model was close to the null (adjusted HR = 0.98, 95% CI: 0.44, 2.20). (**eTable 4**) The estimate from the traditional Cox model was 1.17 (95% CI: 0.87, 1.58).

## DISCUSSION

In summary, regression discontinuity appeared superior to propensity score matching in replication of the known protective association of statins with MI, although precision was limited. Regression discontinuity analysis resulted in relative risk point estimates similar to those from randomized controlled trials in direction and magnitude, whereas matching did not. Meanwhile, propensity matched analyses showed an increased risk of MI among statin users, likely due to confounding by indication. While both regression discontinuity design and propensity score matching achieved good balance of measured confounders, we found that only regression discontinuity methods achieved estimates nearing those from trials, which are in the range of a relative risk of 0.7 to 0.8.^2,3,33^ The estimate for the association of statin use and injuries from motor vehicle accidents, as a negative control outcome, was close to the null in a regression discontinuity model. In sum, the performance of the regression discontinuity design suggests that this approach may account for bias from measured as well as unmeasured confounders.

Our study builds on prior methodologic investigations that have aimed to estimate the causal effect of statin use in the THIN database. Geneletti and O’Keeffe and others have previously validated the regression discontinuity design to estimate causal effects when applied to estimate the effect of statins on low density lipoprotein cholesterol concentration for people around the 20% CVD risk threshold in the THIN database.^34,35^ They demonstrated a protective effect of statins on low density lipoprotein cholesterol using regression discontinuity estimates of the average treatment effect, although the effect size was smaller than observed in randomized controlled trials. Using simulation studies, they showed that regression discontinuity estimators were attenuated towards the null in simulations with high unmeasured confounding and non-compliance with the guideline. Our investigation extends this work by evaluating two hard outcomes, MI and motor vehicle accidents, and comparing the findings to a propensity score matched analysis. Although we do not know the true confounding structure in our data, our estimates likely represent an underestimation of the treatment effects of statin use due to incomplete compliance.

A major challenge was that event rates were lower than in trials, which often by design over-sample for high-risk participants. Thus, even with a large sample size, we had a modest number of events and precision of the estimates was limited. Additionally, we used a sharp regression discontinuity design, which assumes that those above the cutoff initiated statins and those below the cutoff did not – as in an intent-to-treat analysis. However, we observed empirically that many deviated from this rule. A fuzzy RD design would relax this assumption allowing for non-compliance. However, methods that incorporates a fuzzy regression discontinuity design into a Cox model are not available, and this is an ongoing area of methodologic development. Moreover, adherence to statin prescriptions in real world settings is moderate and may wane over time.^36-38^ This could explain the attenuation towards the null of the MI effect estimate from the regression discontinuity models within 10 years vs. 5 years of follow up.

Our study provides a real-world application of regression discontinuity estimation leveraging a national guideline on wider statins use in the UK. The latter creates the setting of a natural experiment in which the probability of treatment around the cutoff can be regarded as an exogenous variation in treatment status. Furthermore, the implementation of the guideline provides a unique advantage to estimating causality relative to the US, which does not have a national health care system and is subject to more heterogeneity in practice at the health care system, clinic, and provider level. However, our study also has limitations that should be considered. Electronic health records are subject to misclassification bias. Although previous studies have demonstrated a high degree of validity in THIN data, ^19-21^ there may still be residual measurement error. Because we used electronic health record data, we had no individual-level information on socioeconomic status. Instead, we used the Townsend deprivation score, which is an area-level score indicated socioeconomic status based on Census data. However, in prior work, the Townsend score has been shown to correlate strongly with individual-level SES, including education.^39^ Additionally, this study was conducted in the UK and may not generalize to other populations. However, we have no reason to believe that the biological effects of statins would vary by population or geography.^33,40-43^

In summary, regression discontinuity appears to be a promising approach to control for measured and unmeasured confounding in observational studies. This head-to-head comparison approach will need to be replicated in other cohorts and with different treatments and outcomes before it can become the preferred approach. Importantly, by replicating the statin-MI trial estimates using RD, our findings suggest that, when used appropriately, quasi-experimental methods can expand the scope of clinical investigations, making it feasible to estimate causal effects in observational data.

## Data Availability

The data utilized for this study are available from The Health Improvement Network (THIN; https://www.the-health-improvement-network.com/en/). However, restrictions apply to the availability of this data, and so are not publicly available. However, data are available upon reasonable request and with permission of THIN.

## Funding

This work was funded by R56-AG061177 from the National Institute on Aging.

## Supplemental Material

**eTable 1:** Summary of CVD Risk Scores by Type

|  | N | Mean | SD |
| --- | --- | --- | --- |
| 1991 Framingham 10-year | 40,259 | 23.5 | 10.60 |
| QRISK | 2,868 | 22.4 | 9.69 |
| QRISK2 | 5,858 | 21.1 | 10.0 |
| Multiple Scores | 257 | 22.7 | 9.08 |

**eTable 2:** Adherence to statin treatment below and above the 20% CVD risk score cutoff

|  | Below 20% | Above 20% |
| --- | --- | --- |
| Total | 21,326 | 27,916 |
| Not Prescribed Statins | **18,788 (88.1%)** | 18,047 (64.6%) |
| Prescribed Statins | 2,538 (11.9%) | **9,869 (35.4%)** |

**eTable 3:**
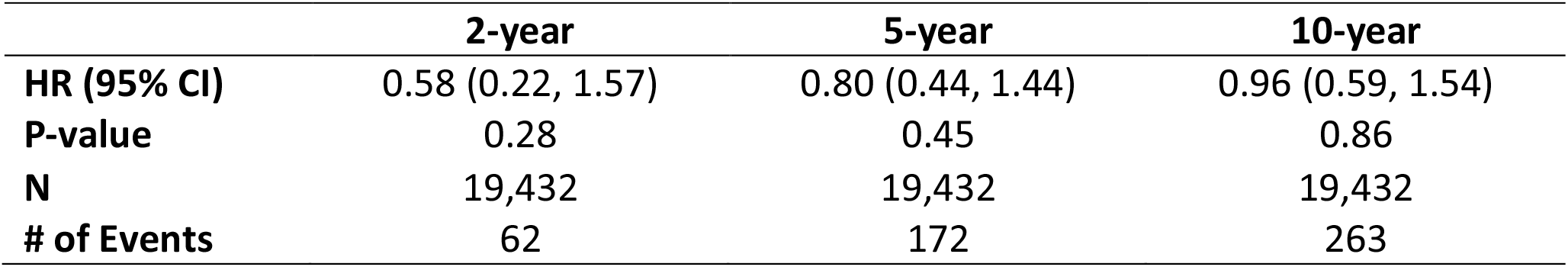
Regression discontinuity estimates of the association of statins use (2008-2011) and incident MI over 2, 5, and 10 years

**eTable 4:** A comparison of estimates of the association of statins and motor vehicle accidents over 10 years, based on regression discontinuity and Cox models

| Model | N | HR | Confidence Interval | p-value |
| --- | --- | --- | --- | --- |
| **Regression Discontinuity Model\*** |  |  |  |  |
| Sex, Age adjusted | 19,432 | 0.97 | 0.43, 2.17 | 0.93 |
| Fully<sup>†</sup> adjusted | 19,432 | 0.98 | 0.44, 2.20 | 0.96 |
| **Traditional Cox Model (Full cohort)** |  |  |  |  |
| Sex, Age adjusted | 49,242 | 1.18 | 0.88, 1.59 | 0.27 |
| Fully<sup>†</sup> adjusted | 49,242 | 1.17 | 0.87, 1.58 | 0.30 |

\*This sample was restricted to those within the bandwidth of 5.0
<sup>†</sup> Adjusted for age, sex, marital status, Townsend score, smoking, hypertension; inclusion of diabetes, physical activity, or BMI resulted in lack of convergence.

**eFigure 1:**
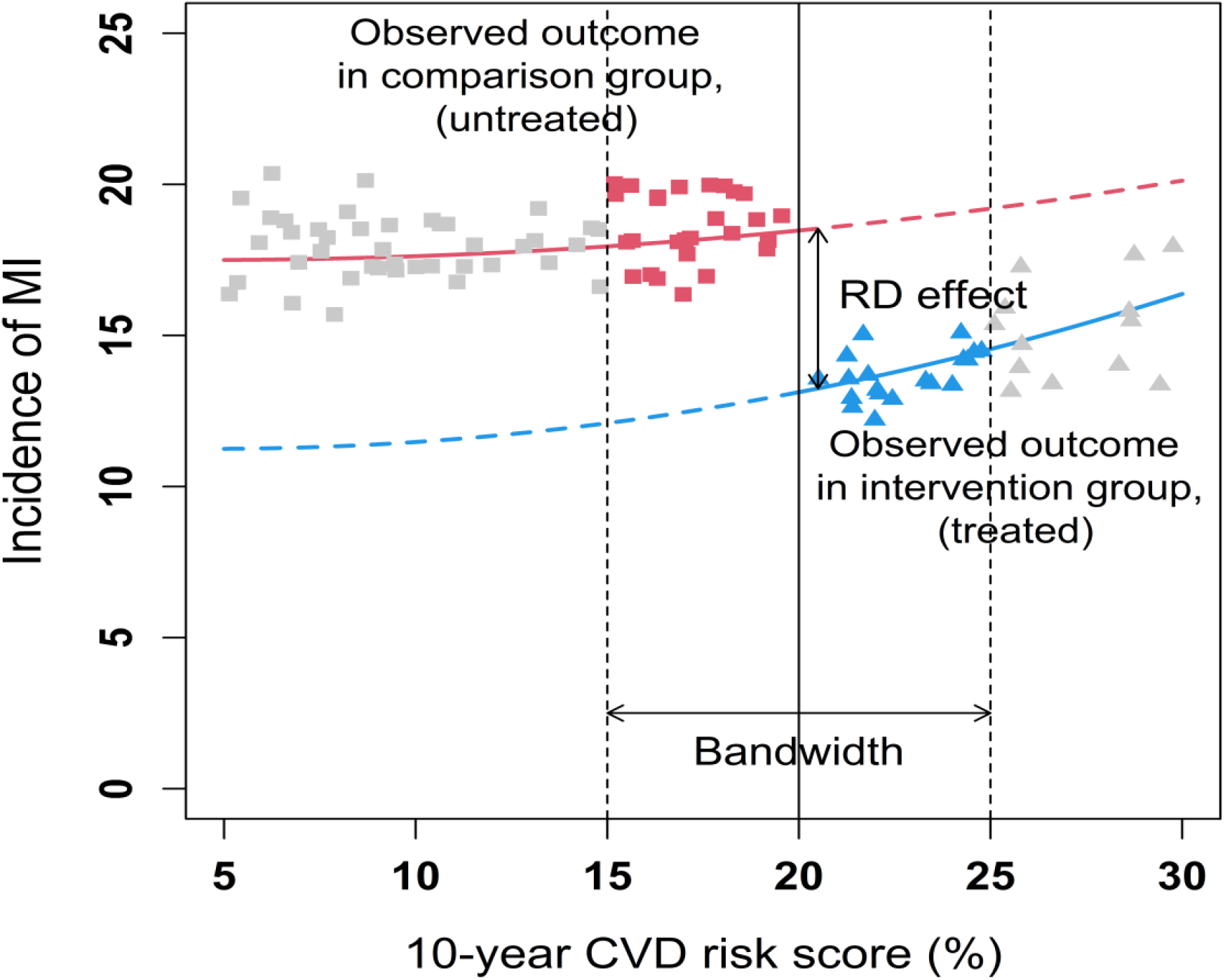
Illustration of RDD design and estimation of statin treatment effects on incident MI This figure is a hypothetical illustration of an RD design to estimate the treatment effect of statin medication on dementia risk. The x-axis displays the *<u>instrument</u>*, which is 10-year CVD risk score, together with the cutoff (e.g. of 20%), set by the guideline. Those with a 10-year CVD risk score ≥ 20% will be more likely to be prescribed statin (i.e., Treated = 1, solid **blue** line). Statin will be less likely to be prescribed in patients with a 10-year CVD risk score <20% (i.e., Treated = 0, solid **red** line). The RD effect, which equals the vertical distance between the blue and red lines at the cutoff, is the **causal treatment effect of interest**.

**eFigure 2.**
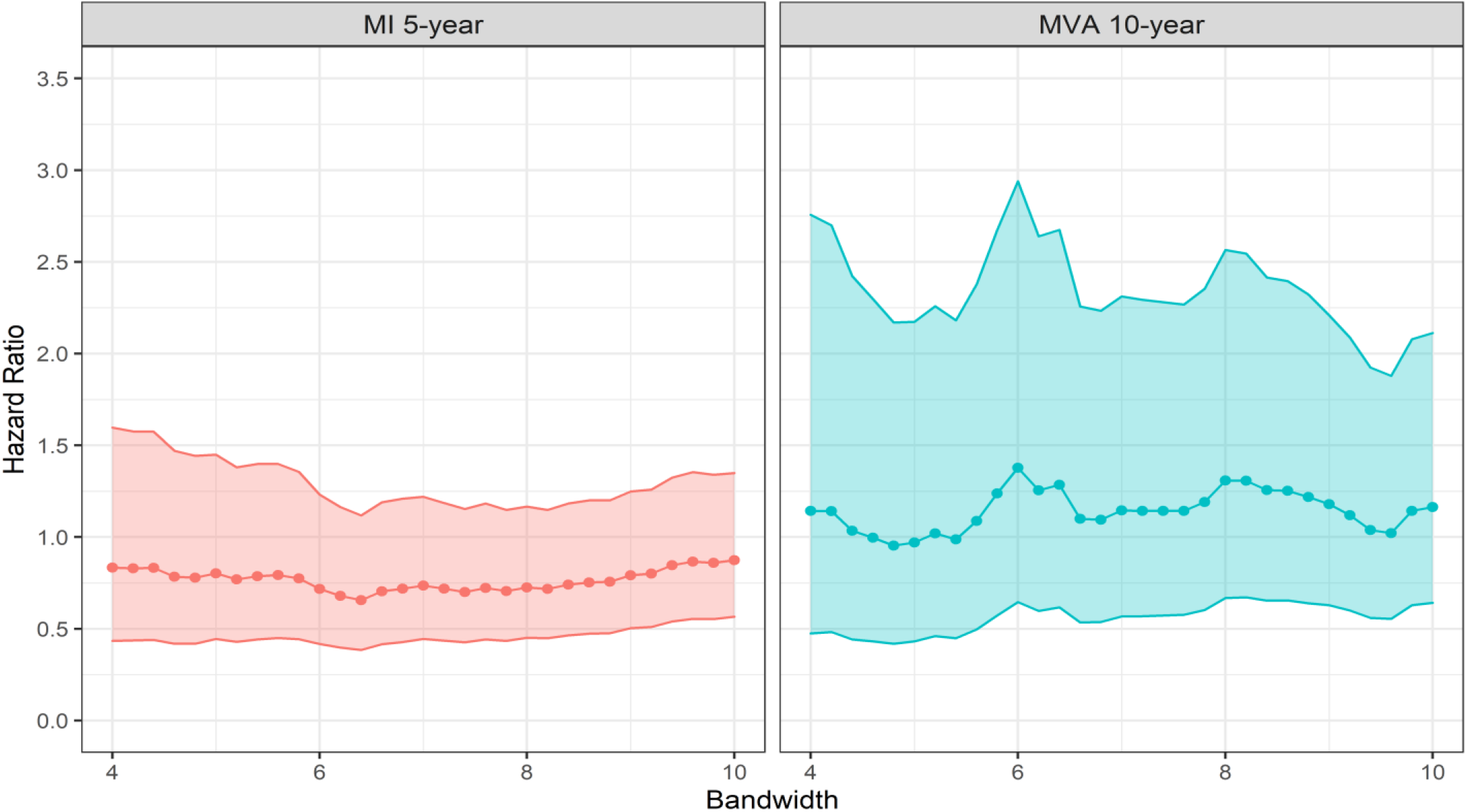
Bandwidth calibration illustrates bandwidth on X-axis and HR for 5-year MI (left) and 10-year MVA (right)

**eFigure 3:**
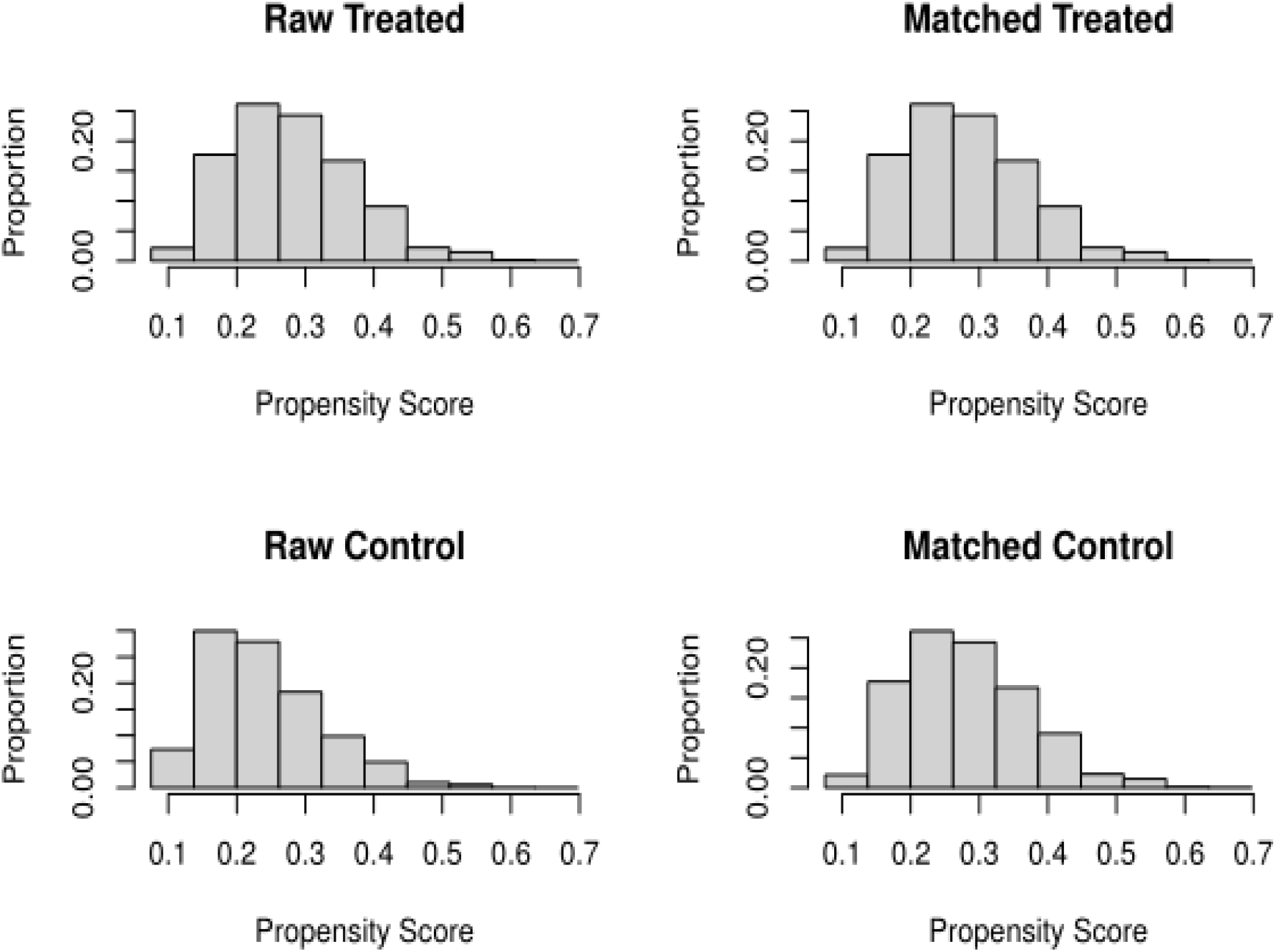
Histograms of propensity scores for statin treatment, before and after matching

## Notes

### Competing Interest Statement

The authors have declared no competing interest.

### Author Declarations

IRB of Columbia University gave ethical approval for this work

### Summary of Updates

Title revised; Abstract revised; Figure 2, eFigure 1, and eFigure 2 revised; further details provided in Introduction and Methods sections, leading to some updates to the Results; further Discussion points added

## References

1. Cholesterol Treatment Trialists C, Fulcher J, O’Connell R, et al. Efficacy and safety of LDL-lowering therapy among men and women: meta-analysis of individual data from 174,000 participants in 27 randomised trials. Lancet. 2015;385(9976):1397–1405.

2. Shepherd J, Blauw GJ, Murphy MB, et al. Pravastatin in elderly individuals at risk of vascular disease (PROSPER): a randomised controlled trial. Lancet. 2002;360(9346):1623–1630.

3. Han BH, Sutin D, Williamson JD, et al. Effect of Statin Treatment vs Usual Care on Primary Cardiovascular Prevention Among Older Adults: The ALLHAT-LLT Randomized Clinical Trial. JAMA Intern Med. 2017;177(7):955–965.

4. Hippisley-Cox J, Coupland C, Vinogradova Y, Robson J, Brindle P. Performance of the QRISK cardiovascular risk prediction algorithm in an independent UK sample of patients from general practice: a validation study. Heart. 2008;94(1):34–39.

5. Venkataramani AS, Bor J, Jena AB. Regression discontinuity designs in healthcare research. BMJ. 2016;352:i1216.

6. Wallace J, Jiang K, Goldsmith-Pinkham P, Song Z. Changes in Racial and Ethnic Disparities in Access to Care and Health Among US Adults at Age 65 Years. JAMA Intern Med. 2021;181(9):1207–1215.

7. Fukuma S, Iizuka T, Ikenoue T, Tsugawa Y. Association of the National Health Guidance Intervention for Obesity and Cardiovascular Risks With Health Outcomes Among Japanese Men. JAMA Intern Med. 2020;180(12):1630–1637.

8. Maciejewski ML, Basu A. Regression Discontinuity Design. JAMA. 2020;324(4):381–382.

9. Desai S, McWilliams JM. Consequences of the 340B Drug Pricing Program. N Engl J Med. 2018;378(6):539–548.

10. Cattaneo MD, Titiunik R. Regression Discontinuity Designs. Annual Review of Economics. Volume 14, 2022. Forthcoming.

11. Hahn J, P. Todd, Klaauw Wvd. Identification and Estimation of Treatment Effects with a Regression-Discontinuity Design. Econometrica. 2001;69(1):201–209.

12. Lee DS, Lemieux T. Regression Discontinuity Designs in Economics Journal of Economic Literature. 2010;48:281–355.

13. Thistlethwaite DL, Campbell DT. Regression-Discontinuity Analysis: An Alternative to the Ex-Post Facto Experiment. Journal of Educational Psychology. 1960;51(6):309–317.

14. Rosenbaum PR, Rubin DB. The Central Role of the Propensity Score in Observational Studies for Causal Effects. Biometrika. 1983;70(1):41–55.

15. Cattaneo MD, Idrobo N, R T. A practical introduction to regression discontinuity designs: Foundations. Cambridge University Press; 2019 Nov.

16. Blak BT, Thompson M, Dattani H, Bourke A. Generalisability of The Health Improvement Network (THIN) database: demographics, chronic disease prevalence and mortality rates. Inform Prim Care. 2011;19(4):251–255.

17. Danaei G, Garcia Rodriguez LA, Fernandez Cantero O, Hernan MA. Statins and risk of diabetes: an analysis of electronic medical records to evaluate possible bias due to differential survival. Diabetes Care. 2013;36(5):1236–1240.

18. Smeeth L, Douglas I, Hall AJ, Hubbard R, Evans S. Effect of statins on a wide range of health outcomes: a cohort study validated by comparison with randomized trials. Br J Clin Pharmacol. 2009;67(1):99–109.

19. Lewis JD, Schinnar R, Bilker WB, Wang X, Strom BL. Validation studies of the health improvement network (THIN) database for pharmacoepidemiology research. Pharmacoepidemiol Drug Saf. 2007;16(4):393–401.

20. Maguire A, Blak BT, Thompson M. The importance of defining periods of complete mortality reporting for research using automated data from primary care. Pharmacoepidemiol Drug Saf. 2009;18(1):76–83.

21. Ruigomez A, Martin-Merino E, Rodriguez LA. Validation of ischemic cerebrovascular diagnoses in the health improvement network (THIN). Pharmacoepidemiol Drug Saf. 2010;19(6):579–585.

22. Bourke A, Dattani H, Robinson M. Feasibility study and methodology to create a quality-evaluated database of primary care data. Inform Prim Care. 2004;12:171–177.

23. Anderson KM, Odell PM, Wilson PW, Kannel WB. Cardiovascular disease risk profiles. Am Heart J. 1991;121(1 Pt 2):293–298.

24. Hippisley-Cox J, Coupland C, Vinogradova Y, Robson J, May M, Brindle P. Derivation and validation of QRISK, a new cardiovascular disease risk score for the United Kingdom: prospective open cohort study. BMJ. 2007;335(7611):136.

25. Hippisley-Cox J, Coupland C, Vinogradova Y, et al. Predicting cardiovascular risk in England and Wales: prospective derivation and validation of QRISK2. BMJ. 2008;336(7659):1475–1482.

26. Garcia Rodriguez LA, Perez Gutthann S. Use of the UK General Practice Research Database for pharmacoepidemiology. Br J Clin Pharmacol. 1998;45(5):419–425.

27. Dave S, Petersen I. Creating medical and drug code lists to identify cases in primary care databases. Pharmacoepidemiol Drug Saf. 2009;18(8):704–707.

28. Chisholm J. The Read clinical classification. BMJ. 1990;300(6732):1092.

29. Calonico S, Cattaneo MD, Titiunik R. Robust Nonparametric Confidence Intervals for Regression-Discontinuity Designs. Econometrica. 2014;82(6):2295–2326.

30. Imbens G, Kalyanaraman K. Optimal Bandwidth Choice for the Regression Discontinuity Estimator. The Review of Economic Studies. 2011;79(3):933–959.

31. Calonico S, Cattaneo M, Titiunik R. Robust Nonparametric Confidence Intervals for Regression-Discontinuity Designs. Econometrica. 2014;82.

32. Calonico S, Cattaneo MD, Farrell MH, Titiunik R. rdrobust: Software for regression-discontinuity designs. Stata J. 2017;17(2):372–404.

33. Cholesterol Treatment Trialists C, Baigent C, Blackwell L, et al. Efficacy and safety of more intensive lowering of LDL cholesterol: a meta-analysis of data from 170,000 participants in 26 randomised trials. Lancet. 2010;376(9753):1670–1681.

34. Geneletti S, O’Keeffe AG, Sharples LD, Richardson S, Baio G. Bayesian regression discontinuity designs: incorporating clinical knowledge in the causal analysis of primary care data. Stat Med. 2015;34(15):2334–2352.

35. O’Keeffe AG, Geneletti S, Baio G, Sharples LD, Nazareth I, Petersen I. Regression discontinuity designs: an approach to the evaluation of treatment efficacy in primary care using observational data. BMJ. 2014;349:g5293.

36. Cohen JD, Brinton EA, Ito MK, Jacobson TA. Understanding Statin Use in America and Gaps in Patient Education (USAGE): an internet-based survey of 10,138 current and former statin users. J Clin Lipidol. 2012;6(3):208–215.

37. Jackevicius CA, Mamdani M, Tu JV. Adherence with statin therapy in elderly patients with and without acute coronary syndromes. JAMA. 2002;288(4):462–467.

38. Lauffenburger JC, Robinson JG, Oramasionwu C, Fang G. Racial/Ethnic and gender gaps in the use of and adherence to evidence-based preventive therapies among elderly Medicare Part D beneficiaries after acute myocardial infarction. Circulation. 2014;129(7):754–763.

39. Adams J, Ryan V, White M. How accurate are Townsend Deprivation Scores as predictors of self-reported health? A comparison with individual level data. J Public Health (Oxf). 2005;27(1):101–106.

40. Yusuf S, Bosch J, Dagenais G, et al. Cholesterol Lowering in Intermediate-Risk Persons without Cardiovascular Disease. N Engl J Med. 2016;374(21):2021–2031.

41. Sattar N, Preiss D, Murray HM, et al. Statins and risk of incident diabetes: a collaborative meta-analysis of randomised statin trials. Lancet. 2010;375(9716):735–742.

42. Ridker PM, Danielson E, Fonseca FA, et al. Rosuvastatin to prevent vascular events in men and women with elevated C-reactive protein. N Engl J Med. 2008;359(21):2195–2207.

43. Officers A, Coordinators for the ACRGTA, Lipid-Lowering Treatment to Prevent Heart Attack T. Major outcomes in moderately hypercholesterolemic, hypertensive patients randomized to pravastatin vs usual care: The Antihypertensive and Lipid-Lowering Treatment to Prevent Heart Attack Trial (ALLHAT-LLT). JAMA. 2002;288(23):2998–3007.

